# Tackling the primary healthcare workforce crisis: time to talk about health systems and governance. A comparative assessment of the European region

**DOI:** 10.1101/2024.05.24.24307895

**Authors:** E Kuhlmann, M Falkenbach, MG Brinzac, T Correia, M Panagioti, B Rechel, A Sagan, M Santric-Milicevic, M-I Ungureanu, I Wallenburg, V Burau

## Abstract

**Background:** Primary healthcare has emerged as a powerful global concept, but little attention has been directed towards the pivotal role of the healthcare workforce and the diverse institutional setting in which they work. This study aims to bridge the gap between the primary healthcare policy and the ongoing healthcare workforce crisis debate by introducing a health system and governance approach to identify transformative capacities in health system contexts.

**Methods:** A qualitative comparative methodology was employed, and a rapid assessment of the primary healthcare workforce was conducted across nine countries: Denmark, Germany, Kazakhstan, Netherlands, Portugal, Romania, Serbia, Switzerland, and the United Kingdom/ England.

**Results:** Our findings reveal both convergence and pronounced diversity across the healthcare systems, with none fully aligning with the ideal attributes of primary healthcare suggested by WHO. However, across all categories, Denmark, the Netherlands, and to a lesser extent Kazakhstan, depict closer alignment to this model than the other countries. Workforce composition and skill-mix vary strongly, while disparities persist in education and data availability, particularly within Social Health Insurance systems. Policy responses and interventions span governance, organisational, and professional realms, although with weaknesses in the implementation of policies and a systematic lack of data and evaluation. The WHO primary healthcare model only marginally informs policy decisions, with the exception being in Kazakhstan.

**Conclusion:** We conclude that aligning primary healthcare and workforce considerations within the broader health system context may help move the debate forward and build governance capacities to improve resilience in both areas.

## Background

Primary healthcare (PHC) has emerged as a powerful global concept aiming to improve health outcomes, enhancing health system efficiency, resilience, and equity. It has inspired many policy changes, advocating for its prioritisation on the global health agenda [1–12]. Within the European region, PHC has been the focal point of discussions in various meetings of the World Health Organisation (WHO) signalling a collective commitment to action [13–14]. These discussions have been reinforced by advancements in data and research including on the healthcare workforce (HCWF) [6, 15–20], providing evidence of PHC’s efficiency, specifically during crisis conditions [9, 21–25].

PHC ‘stands as the principal interface between the health system and communities’ [26, website], embodying the convergence of public health and medical care [7, 27]. Defined as a whole-of-society approach, it is based on multisectoral and inclusive policies supporting ‘first-contact, accessible, continued, comprehensive and coordinated patient-focused care’ [7, see also, 4, 26]. The PHC workforce ‘includes all occupations engaged in health promotion, disease prevention, treatment, rehabilitation, and palliative care services, as well as addressing the social determinants of health inclusive of caregivers and volunteers’ [28]. Multidisciplinary teams comprising of various healthcare professionals play a key role in delivering effective PHC services [19, 28].

Although significant progress has been made in advancing PHC [4, 8, 13, 29], some gaps persist, particularly surrounding the integration of PHC and HCWF debates and its prioritisation within national health agendas [30]. The disconnect between PHC implementation and HCWF exasperates the workforce crisis and obstructs effective service delivery. Recent WHO efforts [26] have underscored the challenges in aligning the global PHC-oriented model with national health systems [13], highlighting the need for tailored approaches to fulfil diverse needs and contexts. Others have addressed the ‘layered’ nature of the PHC workforce crisis and emphasised that the ‘causes at the heart of such crisis, and their patterns and implications differ across the very diverse European region’ [30]. Longing for a broad and inclusive ‘one-size-fits-all’ PHC model may risk obscuring these layers and diverse conditions and institutional prerequisites, including powerful professional stakeholders and interests, necessary for effective policy recommendations [12, 20, 31–34].

PHC’s far-reaching promises of universal health coverage (UHC) [4, 29, 35] and equity [4, 29, 36] strengthen the appeal of uniform concepts silencing the critical debate and eventual emerging controversies pertaining to strategies, actors and future directions [12, 20, 32, 37–38]. Crucially, governance largely remains a black box, hiding the systematic analysis of transformative capacities and making PHC workforce governance poorly prepared for the implementation challenges embedded in politics, policy, and the powers of stakeholders within health systems.

The HCWF crisis poses significant challenges to PHC provision, yet the strategic importance of the HCWF as the backbone of every PHC system and the transformative role of HCWs remain understudied and underappreciated. HCWs are largely reduced to ‘operational’ dimensions [6, Figure 1], ignoring their role as agents and professional policy actors [39]. This oversight inflames the burden on individual HCWs [40–44] hindering recruitment, retention, and workforce resilience [45–46]. Addressing these challenges will require an increase in knowledge exchange between countries looking for evidence-based good practice strategies.

Despite the importance of comparative PHC workforce research, it remains underdeveloped with broad and diverse definitions (for an overview, see [26, Table 3.1]) and recommendations [6, 19, 28, 47] hindering empirical operationalisation. Key areas, such as the HCWF and composition and functioning of multidisciplinary teams [48, 49] and the role of community health workers [22, 28, 50], require further examination to effectively inform policy and research moving forward. The lack of PHC-disaggregated workforce data and monitoring systems in most countries, except for physicians [16, 18] further complicates efforts to assess PHC performance and identify areas for improvement [48, 51–52].

Our comparative assessment aims to bridge the gap between the PHC policy debate and the HCWF crisis debate by introducing a health system and governance approach to identify transformative capacities in health system contexts and contribute new knowledge that may help respond effectively to the HCWF crisis.

## Methods

We employed a qualitative comparative methodology, which is explorative and informed by health systems and governance theories [11, 53–55]. Drawing upon insights from previous research [39, 41], our study aimed to investigate the complex interplay between PHC workforce dynamics and health system governance within the WHO European region. A rapid assessment of the PHC workforce was performed based on a case study design and expert information.

### The country sample

The country sample includes nine countries: Denmark, Germany, Kazakhstan, Netherlands, Portugal (excluding the Azores and Madeira), Romania, Serbia, Switzerland, and the United Kingdom with a focus on England. The selection aimed to capture a diverse array of healthcare systems including the National Health Service (NHS)/Beveridge, Social Health Insurance (SHI)/Bismarckian, and emergent (post-communist) SHI systems in Central Eastern and Eastern Europe, while also accounting for diversity within ideal-type systems. By going beyond typological categorisations, our study embraced diversity allowing for the examination of various contextual factors to gain deeper insights into the PHC workforce crisis and inform actionable interventions [39, 41, 53–54]. Across countries, various terminologies exist to describe PHC and its practitioners. In this study, we use PHC, ambulatory general care and family care, as well as GP and family physician synonymously. Table 1 provides an overview of the country sample, considering different types of healthcare systems, PHC models, labour market conditions and workforce compositions, and geographical variation [16, 56, (Denmark) 57, (Germany) 58, (Kazakhstan) 59-62, (Netherlands) 63, (Portugal) 64, (Romania) 65, (Serbia) 66-68, (Switzerland) 69, (United Kingdom/England) 70].

**Table 1.**
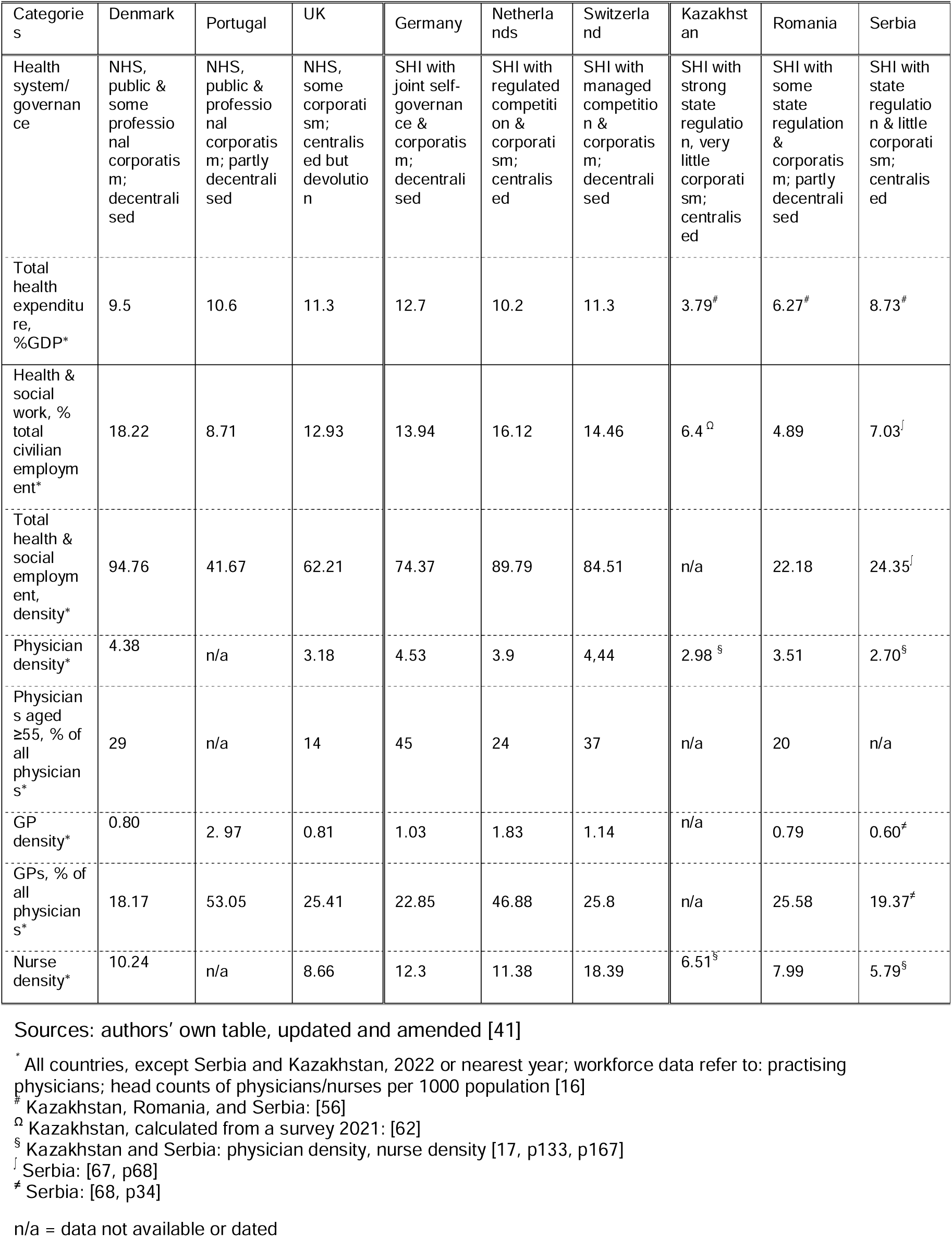
The country sample: health systems and healthcare workforce figures.

| Categories | Denmark | Portugal | UK | Germany | Netherlands | Switzerland | Kazakhstan | Romania | Serbia |
| --- | --- | --- | --- | --- | --- | --- | --- | --- | --- |
| Health system/governance | NHS, public & some professional corporatism; decentralised | NHS, public & professional corporatism; partly decentralised | NHS, some corporatism; centralised but devolution | SHI with joint self-governance & corporatism; decentralised | SHI with regulated competition & corporatism; centralised | SHI with managed competition & corporatism; decentralised | SHI with strong state regulation, very little corporatism; centralised | SHI with some state regulation & corporatism; partly decentralised | SHI with state regulation & little corporatism; centralised |
| Total health expenditure, %GDP* | 9.5 | 10.6 | 11.3 | 12.7 | 10.2 | 11.3 | 3.79<sup>#</sup> | 6.27<sup>#</sup> | 8.73<sup>#</sup> |
| Health & social work, % total civilian employment* | 18.22 | 8.71 | 12.93 | 13.94 | 16.12 | 14.46 | 6.4<sup>Ω</sup> | 4.89 | 7.03<sup>†</sup> |
| Total health & social employment, density* | 94.76 | 41.67 | 62.21 | 74.37 | 89.79 | 84.51 | n/a | 22.18 | 24.35<sup>‡</sup> |
| Physician density* | 4.38 | n/a | 3.18 | 4.53 | 3.9 | 4.44 | 2.98<sup>§</sup> | 3.51 | 2.70<sup>§</sup> |
| Physicians aged ≥55, % of all physicians* | 29 | n/a | 14 | 45 | 24 | 37 | n/a | 20 | n/a |
| GP density* | 0.80 | 2.97 | 0.81 | 1.03 | 1.83 | 1.14 | n/a | 0.79 | 0.60<sup>‡</sup> |
| GPs, % of all physicians* | 18.17 | 53.05 | 25.41 | 22.85 | 46.88 | 25.8 | n/a | 25.58 | 19.37<sup>‡</sup> |
| Nurse density* | 10.24 | n/a | 8.66 | 12.3 | 11.38 | 18.39 | 6.51<sup>§</sup> | 7.99 | 5.79<sup>§</sup> |

Sources: authors' own table, updated and amended [41]
\* All countries, except Serbia and Kazakhstan, 2022 or nearest year; workforce data refer to: practising physicians; head counts of physicians/nurses per 1000 population [16]
<sup>#</sup> Kazakhstan, Romania, and Serbia: [56]
<sup>Ω</sup> Kazakhstan, calculated from a survey 2021: [62]
<sup>§</sup> Kazakhstan and Serbia: physician density, nurse density [17, p133, p167]
<sup>†</sup> Serbia: [67, p68]
<sup>‡</sup> Serbia: [68, p34]
n/a = data not available or dated

### Data collection and analysis

Data collection primarily relied on inputs from country experts and secondary sources, such as public statistics, policy documents, and published research (for details, see supplementary material 1). We developed an assessment tool synthesised from the PHC [3, 6–7, 24, 26–27] and HCWF literature [17, 41, 54–55, 71–73]; relevant items were identified and operationalised into a semi-structured matrix (supplementary material 2). This matrix served as a framework for gathering country specific information focusing on recent developments while considering broader institutional and workforce conditions.

A step-by-step explorative team-based approach guided the iterative analysis process. The lead authors produced summaries of the country cases that were then reviewed and revised in consultation with country experts. The iterative process continued until sufficient coherence and clarity were achieved for each country case. From this analysis, three categories (comprising sub-categories) emerged as a framework for a rapid comparative assessment: PHC systems (Table 2), PHC workforce (Table 3), and PHC workforce action taken (Table 4). System types served to structure and proved a framework for organising the complex qualitative data, enabling an exploration of trends and variations across countries.

**Table 2.**
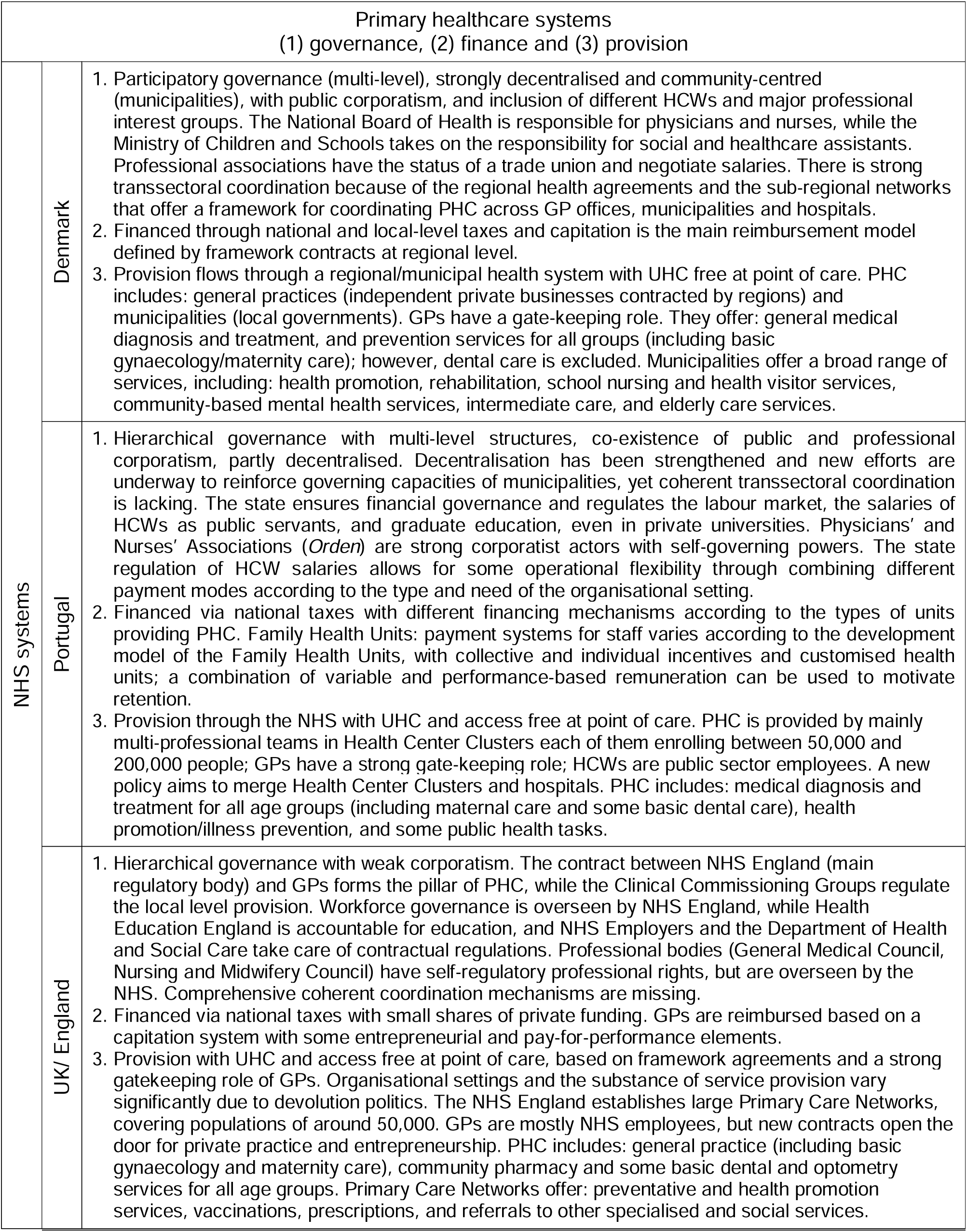
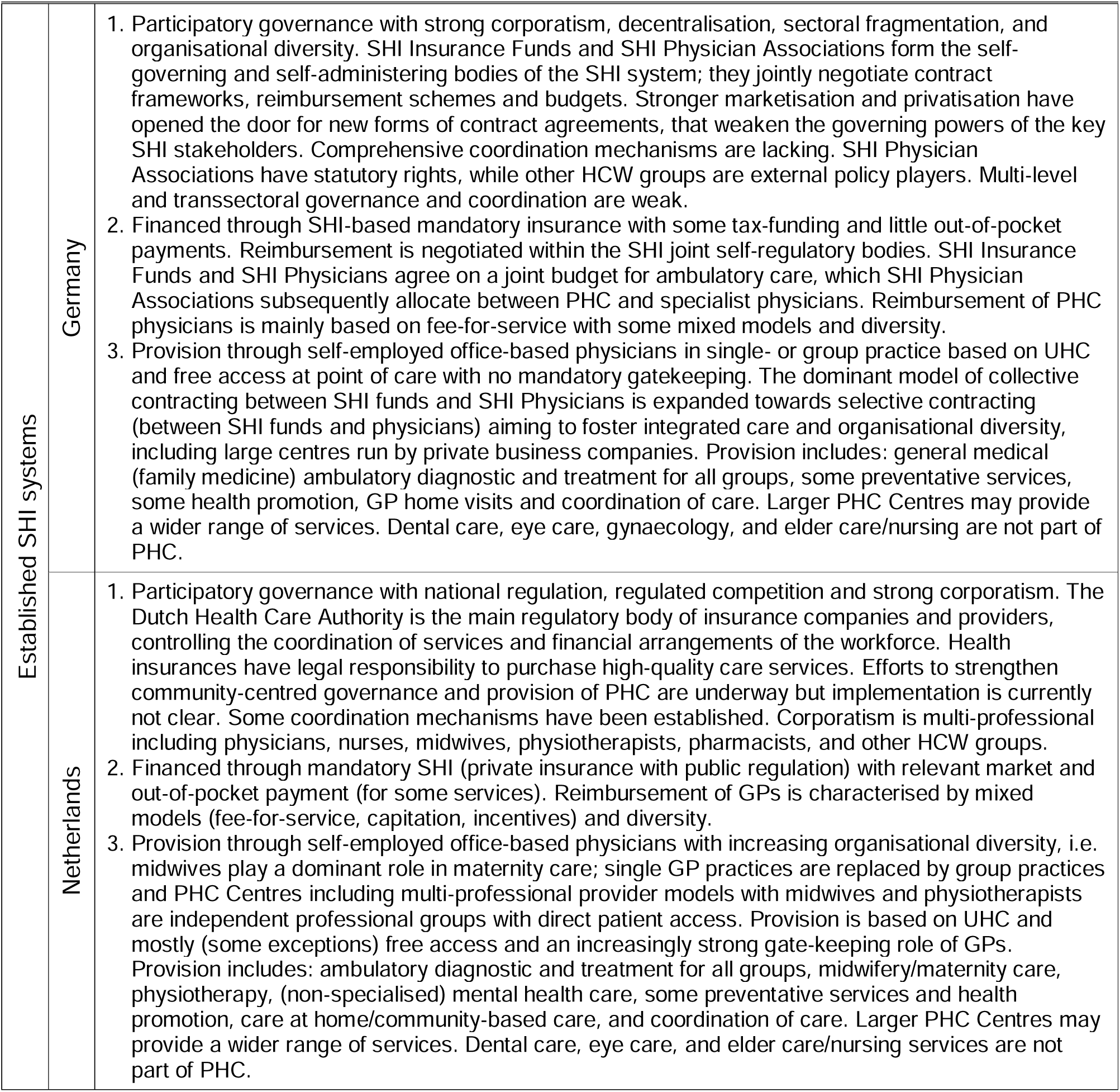
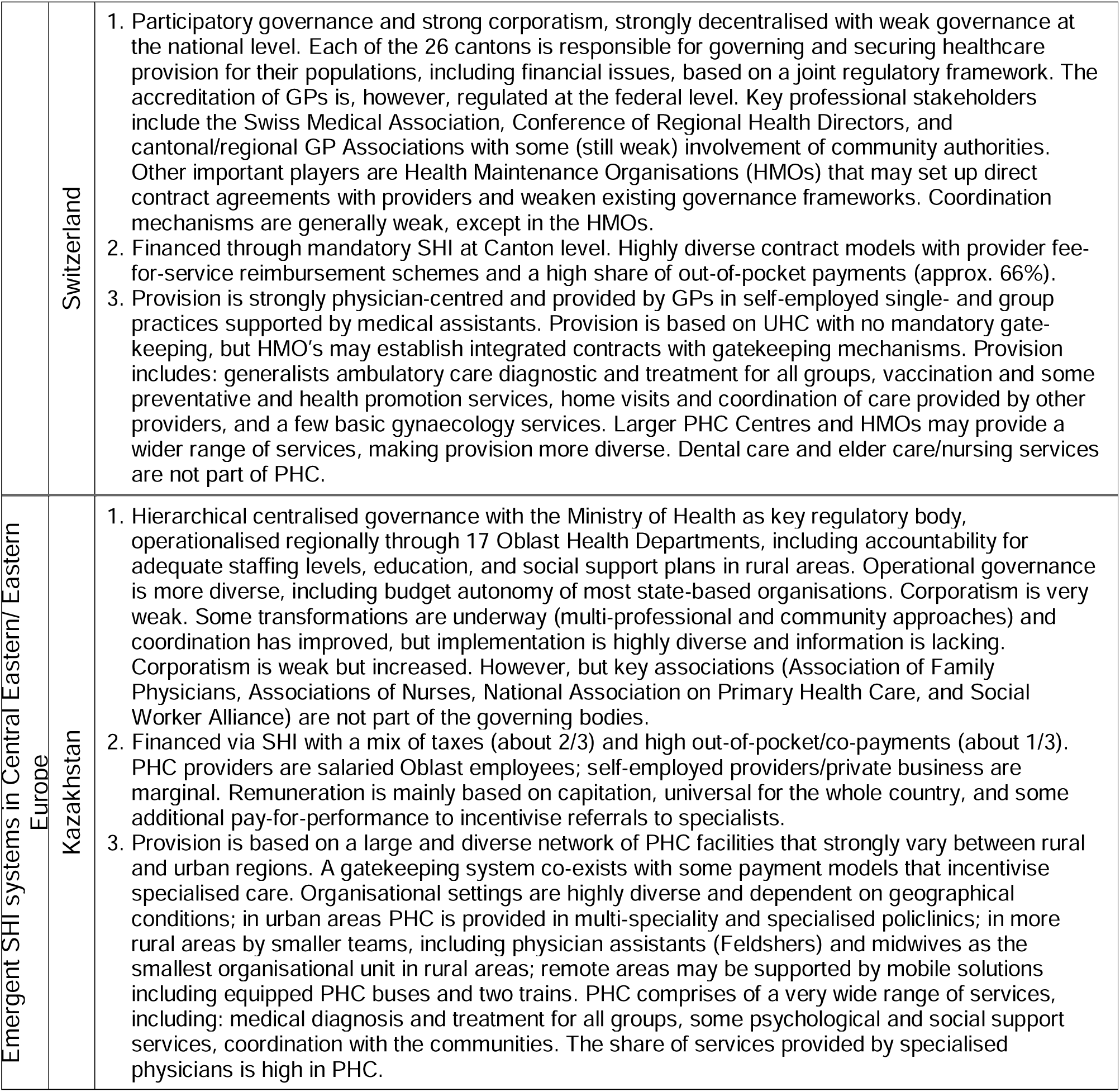
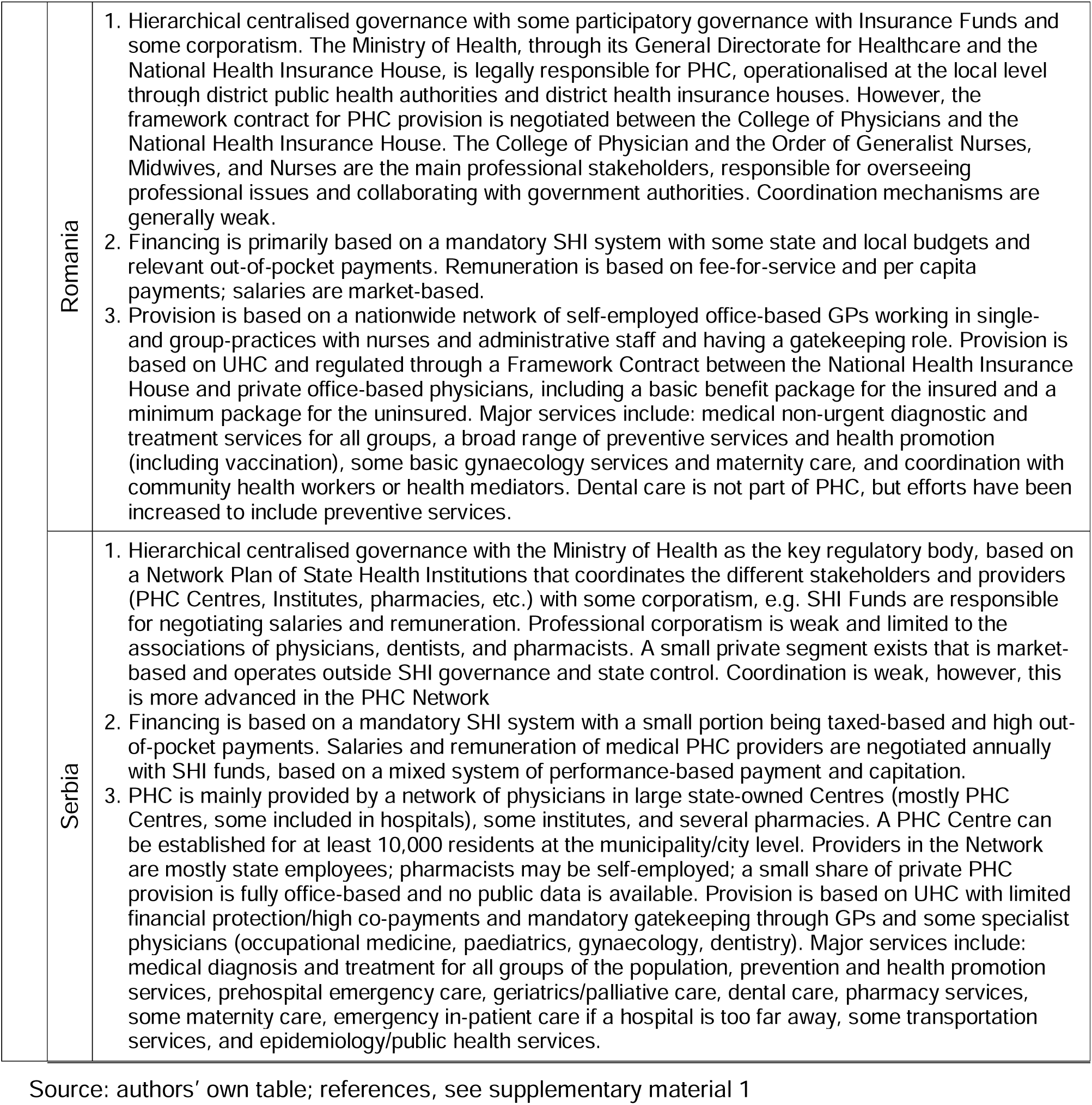
Primary healthcare systems.

**Table 3.**
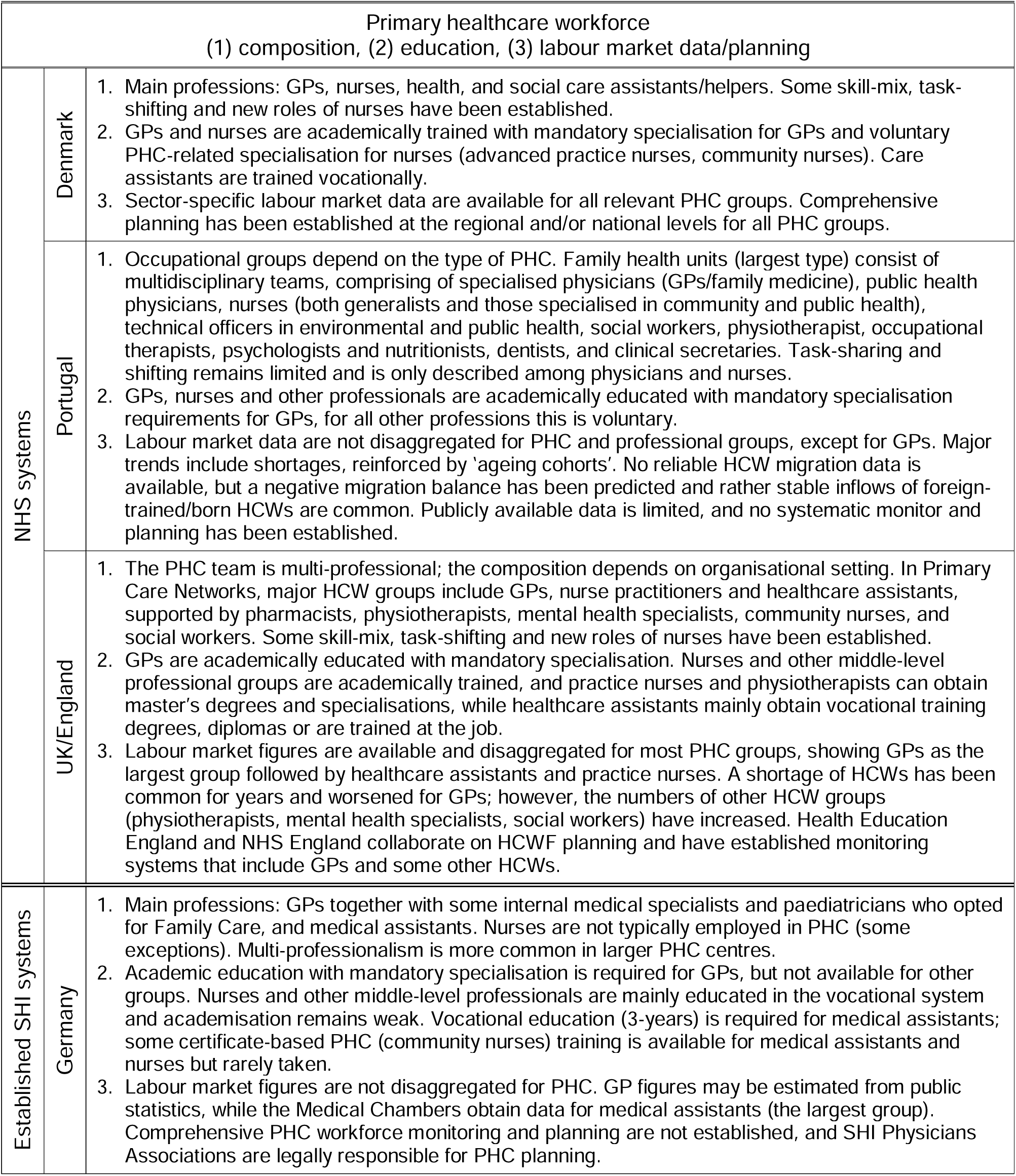
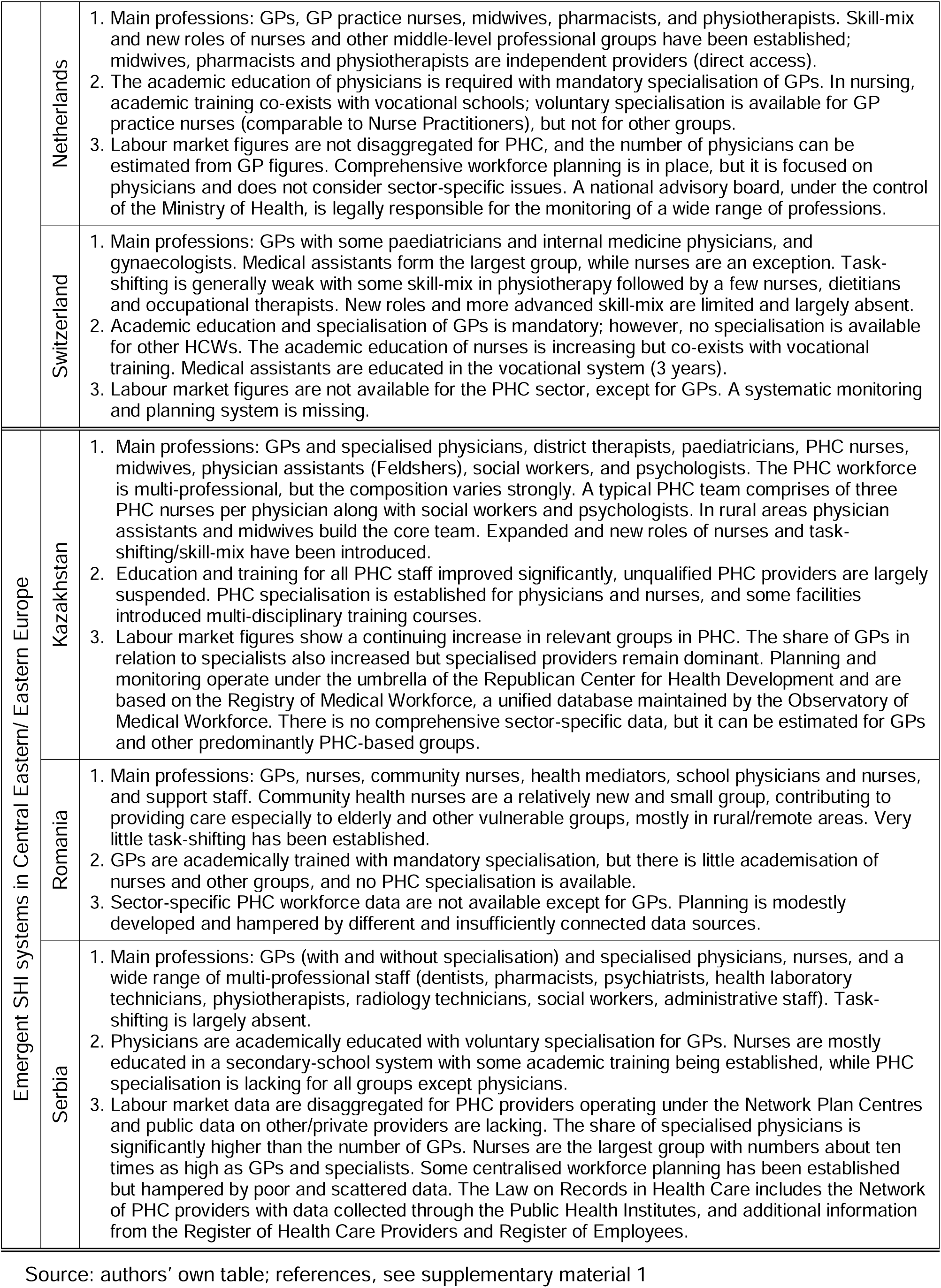
Primary healthcare workforce.

**Table 4.**
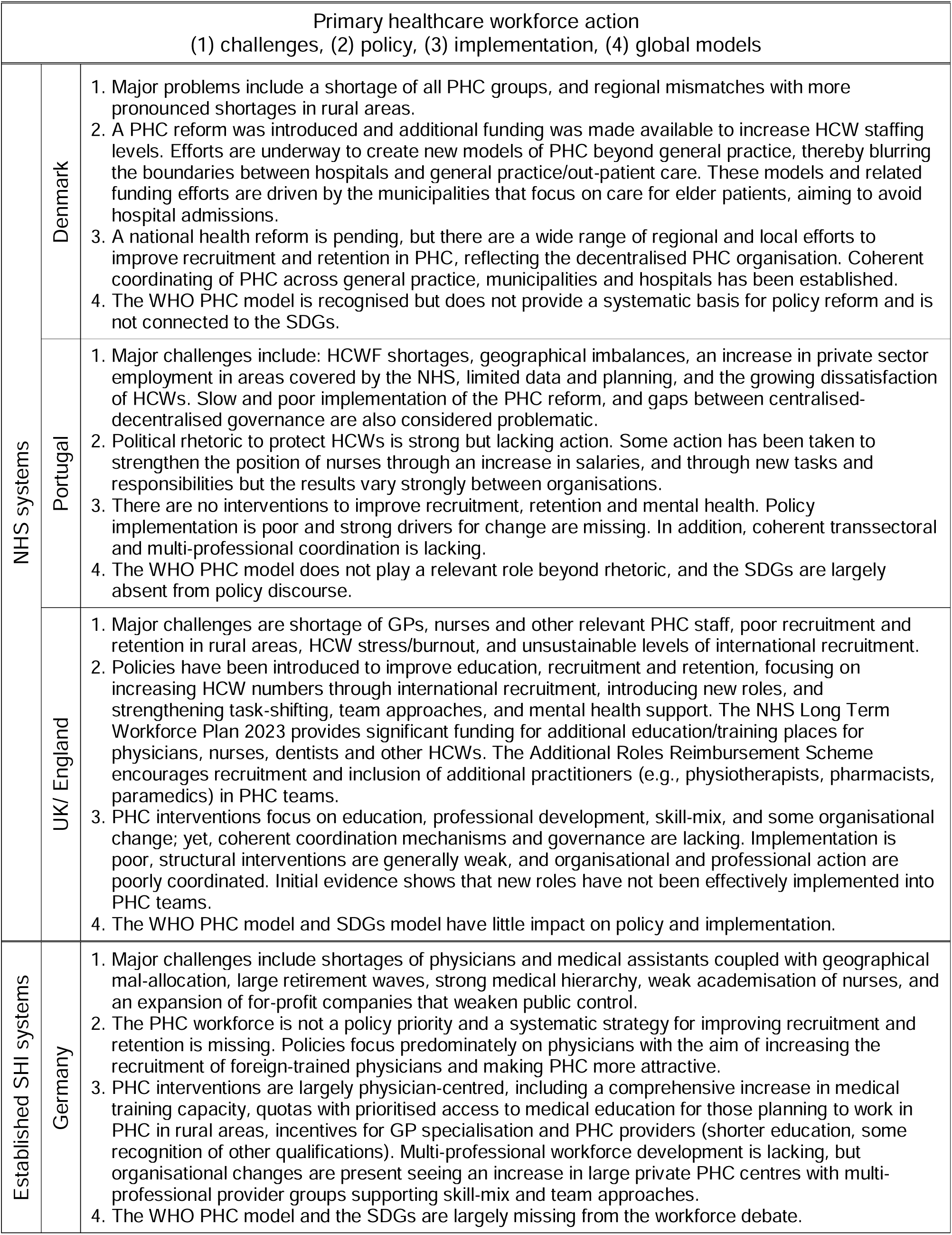
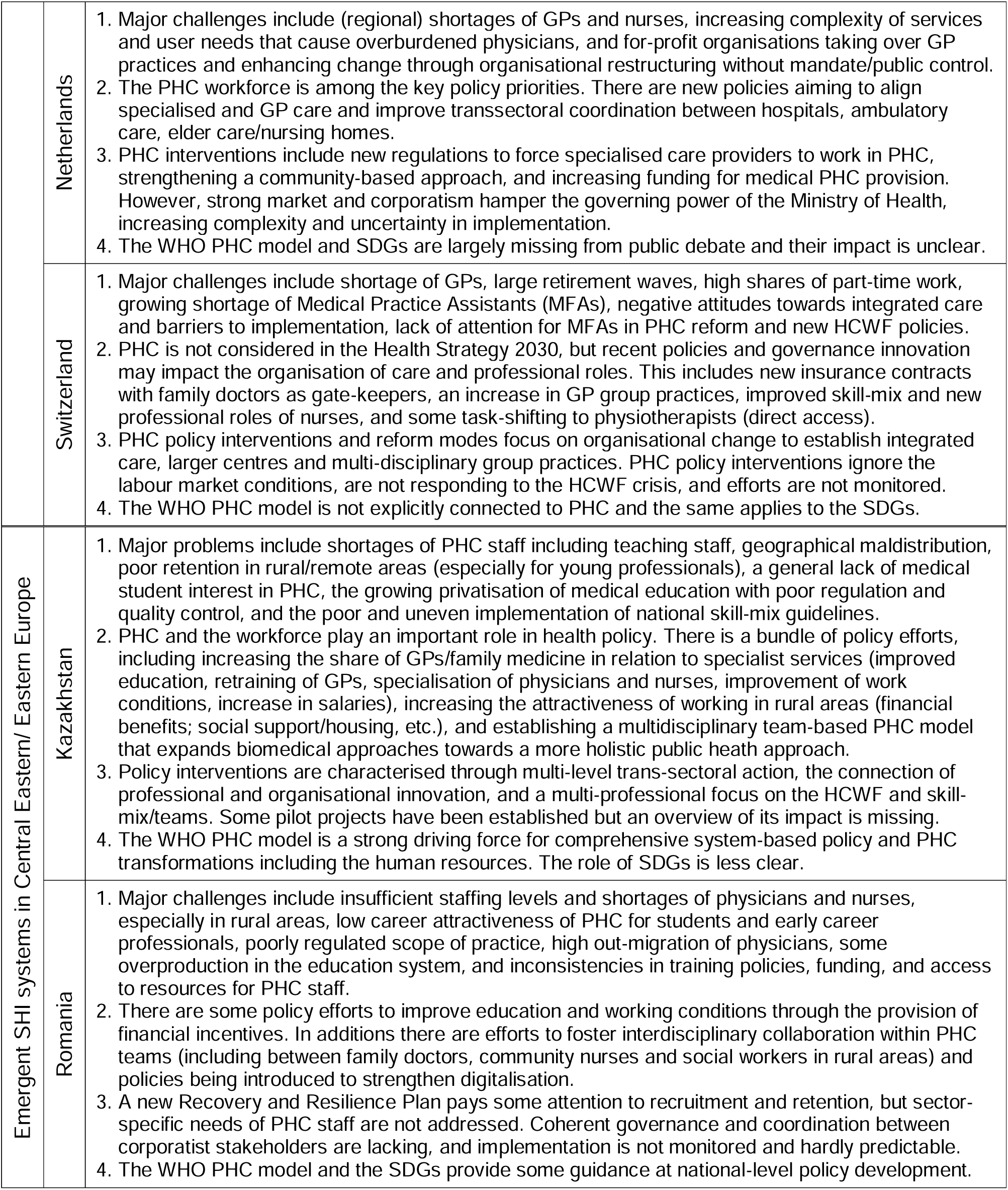
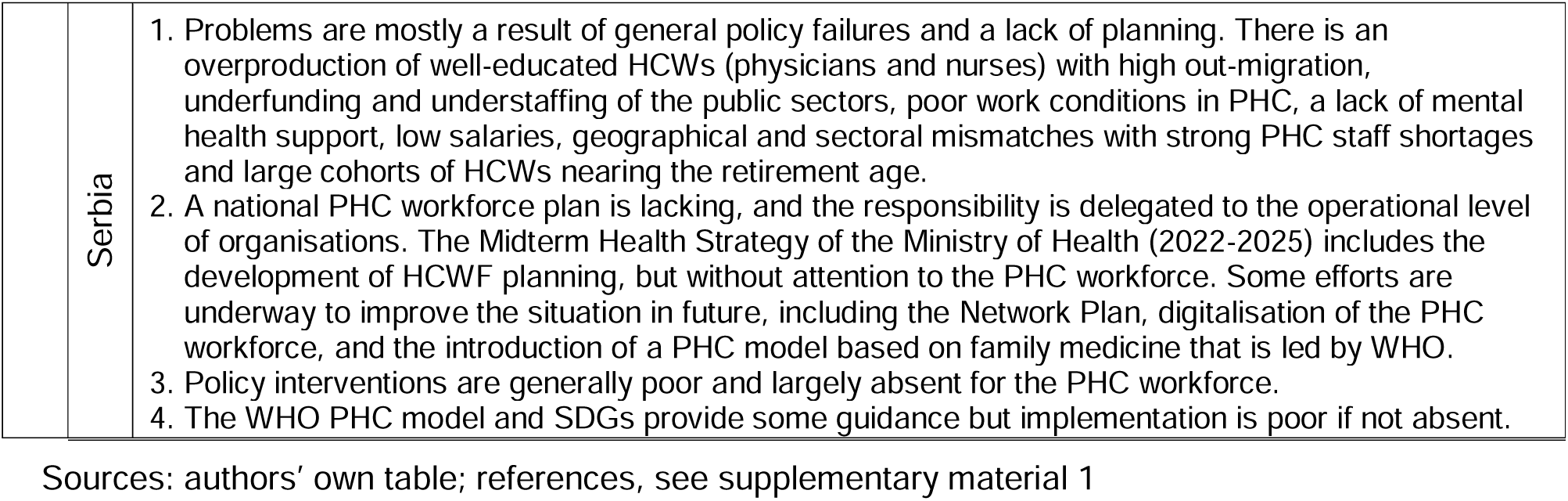
Primary healthcare workforce action.

## Results

### Primary healthcare systems

- NHS systems

The three different NHS systems show some similarities in funding and organisation, yet differences prevail, most strongly in governance. PHC governance ranges from participatory decentralised multi-level governance with comprehensive coordination mechanisms and community-orientated approach in Denmark to more hierarchical forms with weaker coordination in England and Portugal. The inclusion of corporatist actors and self-governing capacities is strong in Denmark and less so in Portugal, and weakest in England. While funding in NHS countries primarily relies on taxes, reimbursement schemes vary, including capitation, pay-for-performance, and mixed incentives. The provision is based on UHC principles with GPs serving as strong gatekeepers and organisational shifts towards larger centres offering comprehensive services for all users. The range of services is generally broad across the countries, including basic maternity care in all countries, and limited dental care available in Portugal and England.

- SHI systems

Historically routed in similar institutions, established SHI systems feature SHI Funds and Physician Associations as key stakeholders and are based on participatory governance and strong corporatism. However, differences have increased in all areas over time, particularly amongst governance structures. Levels of centralisation vary across countries with decentralised governance in Germany and Switzerland and centralised structures in the Netherlands. Coordination mechanisms range from advanced in the Netherlands to weaker in Switzerland and governance transformation efforts differ with some community-centred approaches in the Netherlands, but no substantive changes in the other countries. PHC workforce governance mirrors this complexity with multi-professional bodies and decentralised physicians-centred decision-making with weak coordination in Switzerland and Germany and more advanced efforts in the Netherlands. Funding is mainly based on SHI contributions, with Switzerland and the Netherlands featuring higher shares of private insurances and/or out-of-pocket payments. Fee-for-service dominates reimbursement schemes and provision is largely delivered through contracted private businesses, with a trend towards larger centres and integrated care models. PHC physicians have a gate-keeping role in the Netherlands, while free provider choice prevails in Germany and Switzerland. The range of provisions is similar amongst the three countries and focuses on general medical care with little prevention and promotion services. Specialised care is separate and builds the second pillar of out-patient/ambulatory care. Dental care is not part of PHC, and maternity care is only included in the Netherlands and predominantly led by midwives.

- Emergent SHI systems

Emergent SHI systems exhibit hierarchical governance with varying degrees of centralisation, participatory elements, and stakeholder involvement. Romania focuses on professional corporatism, Serbia emphasises SHI funds, while Kazakhstan demonstrates weak involvement of both. Funding models resemble those of established SHI systems, with SHI contributions supplemented by out-of-pocket payments, although private payments are highest in Kazakhstan. Provision is based on national networks and framework agreements with mandatory gatekeeping and diverse provider models, ranging from mainly self-employed practitioners in Romania to larger state-owned centres in Serbia. Kazakhstan shows stark differences between rural and urban regions and its organisational models include mobile solutions and small units as well as large hospital-based centres. PHC in these countries covers a wide range of basic services for all groups, alongside public health, and some specialised services, although the depth of coverage varies. In Serbia and Kazakhstan, PHC serves as an umbrella for both specialised and generalist providers of out-patient care with specialists taking a powerful position, while GPs are the main PHC providers in Romania.

- Trends across healthcare system types

Overall, although some convergence is discernable, such as more diverse and mixed funding systems and organisational transformations supporting the establishment of larger centres, and increased stakeholder participation in governance, diversity remains pronounced across the PHC systems. None of the countries fully align with the ideal attributes of PHC as highlighted in the WHO framework across all categories, although Denmark, the Netherlands, and to a lesser extent Kazakhstan, come closer to this model than the others.

### The primary care workforce

- NHS systems

Across these systems, there is a largely uniform workforce education and composition characterised by multi-professional teams with physicians and nurses as the largest groups forming the core of PHC. These teams demonstrate some degree of skill-mix, task-shifting and integration of new roles for nurses. Academic education is the dominant model for most groups, including specialised training for nurses. While specialisation is mandatory for GPs in most cases, it remains voluntary for other professions. Sector-disaggregated data for relevant PHC groups is publicly available in all or most NHS countries, except in Portugal, facilitating monitoring and planning efforts. Labour market trends point to GPs shortages and more diverse trends for other groups.

- SHI systems

In established SHI systems, specialised GPs dominate the workforce, although composition varies strongly across countries. The Netherlands stands out for its multi-professional teams including specialised PHC nurses, midwives, physiotherapists, and other providers alongside GPs. Conversely, in Germany and Switzerland, GPs mainly work with medical assistants with task-shifting and skill-mix still in developmental stages. While academic training for GPs is similar across these countries, variations exist for other groups. In the Netherlands, the academisation of a wide range of professional groups is more advanced. For example, the country offers the PHC specialisation of nurses, whereas nurses in Germany and Switzerland lack such opportunities. Sector-disaggregated labour market monitoring is absent across all countries, with only public data available for GPs, and HCWF planning is most comprehensive and advanced in the Netherlands (targeting GPs) and least advanced in Switzerland.

- Emergent SHI systems

In emergent SHI systems, PHC includes a diverse array of professions, with GPs and nurses serving as major groups. Approaches to task-shifting, team-based care and the integration of new roles vary strongly. Kazakhstan is a forerunner in this regard, while Serbia encompasses a traditional hierarchical model with limited task-shifting. Romania finds itself somewhere in the middle of both. Education and specialisation generally improved, with Kazakhstan seeking to up-skill relevant PHC groups and strengthening inter-professional education, while Romania and Serbia continue to focus on physicians. Comprehensive disaggregated PHC data is lacking across all emergent SHI systems except for physicians, posing challenges for workforce planning and evidence-based decision-making.

- Trends across healthcare system types

Overall, this analysis reveals strong differences in the workforce composition. While GPs are central to workforce composition in most countries, Serbia and Kazakhstan feature specialist physicians alongside GPs. Despite ongoing efforts to increase the skill-mix and improve education, disparities persist, particularly in SHI systems (established and emergent) where disaggregated data remains scarce, shaping a workforce planning landscape that is biased towards physicians and hampering evidence generation.

### Primary healthcare workforce action

- NHS systems

Shortage and maldistribution of nurses, physicians, and other relevant PHC groups plague NHS systems, with varying policy approaches across countries. In Denmark, policies focus on structural changes and community-centred approaches supported by bottom-up measures and robust coordination mechanisms. England’s intervention primarily focuses on meso-micro level organisational (some professional) changes, although with poor implementation and ineffective rollouts for new roles despite increased funding for professional development. Portugal shows overall poor interventions and unclear implementation, although some signs of community-centred efforts are emerging. The evaluation of interventions remains sparse across all countries, with Denmark showing more advanced efforts. Neither the WHO PHC-oriented model nor the Sustainable Development Goals (SDGs) have any relevant impact on policies and interventions in these countries.

- Established SHI systems

Shortages of GPs and other PHC staff pose similar challenges in established SHI systems, exacerbated by impending retirement cohorts, particularly in Germany and Switzerland and to a lesser degree in the Netherlands (Table 1). Despite this fact, none of the countries are prioritising the primary HCWF in their policy agendas and no systematic responses have been developed. Interventions mainly focus on organisational changes, although these show a limited focus on governance and weak or altogether absent efforts to improve the skill-mix, task-shifting and the development of new roles, except in the Netherlands. Implementation is hampered in all countries by strong corporatism and market forces, compounded by decentralisation in Germany and Switzerland. As in the NHS systems, the WHO PHC-oriented model and the SDGs have little or no impact on policies and interventions.

- Emergent SHI systems

The emergent SHI systems face similar challenges of workforce shortages and geographical mal-distribution. Serbia and Romania also exhibit a lack of coordination of education and labour markets, high outward-migration rates, and demographic factors all of which do not seem to play a relevant role in Kazakhstan. Policy responses vary strongly between the three countries and are most advanced in Kazakhstan. Here the PHC workforce is prioritised, and multi-professional teams are emphasised. Romania has made some efforts in professional education and team-based approaches, while Serbia lags behind with poor policy efforts. Despite some interventions across all countries, only Kazakhstan has set priorities and taken comprehensive sector-specific action, albeit with challenges pertaining to monitoring and implementation. The WHO PHC-oriented model, and to a lesser degree the SDGs, provide some guidance in all countries, but specifically show strong usage in Kazakhstan where the frameworks drive change in PHC workforce development.

- Trends across healthcare system types

Overall shortages and maldistribution of HCWs are present across all countries. Policy responses and interventions range from governance to organisational, and professional/education measures, although with weaknesses in the implementation of policies and a systematic lack of data and evaluation. The WHO PHC-oriented model and the SDGs only marginally inform policy directions with Kazakhstan standing out for its stronger alignment with these frameworks in comparison to the other countries.

## Discussion

This comparative research provides novel empirical insights that contribute to a deeper understanding of the PHC workforce [30] and prompts critical reflections on the practical implications of the PHC principles put forth in the ‘PHC primer’ [26]. The findings document strong gaps between the global PHC debate and the complex realities of PHC systems and workforce conditions in practice. Our study reveals an absence of typical PHC workforce teams [19, 28, 47–48] and a weak community-orientation across our country samples, with the exception of Denmark (Table 2). This illustrates a substantial implementation gap and accentuates the persisting relevance of institutional pre-requisites in shaping PHC delivery [12, 31–33]. Available tools and a one-size-fits-all discourse provide little opportunity to systematically address the implementation challenges, as diverse system conditions and governance arrangements remain largely invisible. The same holds true when thinking about potential windows of opportunity for transformative policies.

By advocating for a health system approach and shifting the debate from universal global strategies to the nuanced realities of PHC systems, our research stresses the importance of contextual factors in driving change. The empirical evidence produced in our comparative assessment supports this line of argumentation, revealing that each country has taken some action (Table 4), thereby illuminating transformative policies and related governance capacities. Table 5 provides an overview of the diverse paths through which the global PHC debate may, or may not, intersect with the realities of healthcare systems, considering both strategic and operational dimensions of governance. Importantly, the summary is neither exclusive nor comprehensive but seeks to show broader emergent trends.

**Table 5.** Transformative policies and governance capacities: major trends.

| System types | Countries | Transformative policies and governance capacities |  |  |
| --- | --- | --- | --- | --- |
|  |  | System level | Organisational level | Professional level |
| NHS systems | Denmark | X | X | X |
|  | Portugal | (X) |  | X |
|  | UK/England | (X) |  | X |
| established SHI systems | Germany |  | X | (X) |
|  | Netherlands | X | X | X |
|  | Switzerland |  | X |  |
| emrgent SHI systems | Kazakhstan | X | X | X |
|  | Romania | X |  | (X) |
|  | Serbia | X | (X) |  |

Source: authors' own table, based on Table 2, Table 3 and Table 4

Our findings reveal the absence of a coherent pattern pertaining to institutional pre-requisites, PHC workforce conditions, and policy interventions across countries. For instance, the NHS system in Denmark relies on GPs working in independent private businesses contracted by regions, but improved transsectoral coordination and began placing greater emphasis on home care (Table 2). In the SHI system in Netherlands, corporatism and private practices co-exist with complex interventions and more transformative governance, including a wide range of professional groups and stronger public/state control (Table 2). On a negative note, high out-of-pocket payments are characteristic of emergent SHI systems, most pronounced in Kazakhstan [61], but also exist in the Portuguese NHS system [64], and to a lesser degree in the SHI system in Switzerland [69].

The research suggests that isolated policy interventions in workforce governance may not achieve sufficient transformative powers in the PHC system at large. For instance, while initiatives like the introduction of new nursing roles in Portugal [74] and incentives for GPs in Germany [75] show promise, they may fall short of achieving systemic change. On the other hand, multi-level governance actions and policy coordination together with community-centred approaches embody stronger transformative potential that may bring the PHC workforce closer to the global model [7, 26]. However, it is precisely these comprehensive policy approaches that often encounter hurdles in their implementation with many countries lacking effective transsectoral and multi-professional coordination mechanisms needed to address the complex challenges facing PHC provision and workforce crisis.

These findings have important implications for the PHC debate and the development of tools and strategies to effectively respond to the ‘layered crisis’ [30] of the PHC workforce. They call for greater attention to healthcare systems and implementation, addressing existing governance gaps across different levels.

### Limitations

Our study has a few important limitations that should be considered. Firstly, we employ a qualitative explorative approach, which helps identify problems and opportunities but does not allow the drawing of quantitative conclusions with regards to the PHC workforce. Secondly, our rapid assessment relies on expert information and secondary sources, and the selected countries are not exhaustive. Although we aimed for diversity by including countries with varied healthcare systems and HCWF compositions (Table 1), it is possible that other relevant items may have been overlooked. Thirdly, our study does not specifically consider gender disparities [76–78] and other inequalities within the PHC workforce, such as the role of migrant HCWs [17, 79–81]. These are important considerations, but data limitations and the scope of our research precluded their examination. Fourth, complexity of PHC systems and the HCWF make interventions and policy implementation at the interface highly complex processes with eventually diverse outcomes. We identified major dimensions of the ‘layered crisis’ [30] and transformative governance capacities but these may not provide a comprehensive picture of the opportunities. Finally, it is important to recognise that our study should be considered a pilot initiative that demonstrates the need for and benefits of in-depth comparative research. Future studies should build upon our methodological approach in order to research the specific aspects of PHC governance, workforce dynamics, and inequalities more comprehensively.

## Conclusions

Our qualitative comparative assessment argues that integrating the debates surrounding PHC and the HCWF crisis while placing greater emphasis on the role of implementation is paramount. By employing a health system and governance approach along with a rapid comparative assessment using qualitative methods and country case studies, we were able to track transformational policies within the broader health system context. Aligning PHC and HCWF discourse and disentangling the layers of the workforce crisis may help move the debate forward and build governance capacities to improve resilience in both areas. Our conceptual framework and empirical findings support the development of evidence-based and context-sensitive policy recommendations.

Key recommendations:

- Recognise the crucial role of the healthcare workforce as the backbone of PHC systems and advocate for coordinated multi-level governance action to support their effectiveness.
- Shift the PHC debate from idealistic attributes to actionable implementation strategies, emphasising the significance of policy dynamics, political contexts, and diverse stakeholder interests.
- Understand the various existing PHC-oriented models and their dynamics to determine the necessary quantity, competencies, and composition of HCWs and how they can be governed effectively to implement PHC.
- Establish a health system and governance approach together with qualitative comparative PHC workforce studies to identify and develop transformational policies within specific contextual settings.
- Prioritise investments in PHC-disaggregated workforce data and monitoring mechanisms to improve evidence-based policymaking and strategic workforce planning.
- Strengthen knowledge exchange and collaboration among international health organisations, governments, and stakeholders to understand and effectively use diverse transformative policies and governance capacities.

## Data Availability

All data generated and analysed during this study are included in this published article and supplementary information files.

## Supplementary Information

The online version contains supplementary material.

Additional file 1: References for the nine country cases

Additional file 2: Matrix

## Acknowledgements

None.

## Authors’ contributions

EK and MF had the idea, designed the study and the methodological framework for the comparative assessment, coordinated the data collection and analysis, and prepared the draft; all authors contributed a country case study; M-GB provided additional expertise on the youth perspective, BR on the CEE region, VB on the methodological approach and comparative findings; all authors commented on the draft and have read and approved the final version.

## This article did not receive any specific funding

## Declarations

## Ethics approval and consent to participate

Not applicable.

## Consent for publication

Not applicable.

## Abbreviations

APN: Advanced Practice Nurse
EU: European Union
HCW: Healthcare workers
HCWF: Healthcare workforce
NHS: National health service
PHC: Primary healthcare
SDGs: Sustainable Development Goals
SHI: Social health insurance
UHC: Universal health coverage
UK: United Kingdom
WHO: World Health Organisation

