## Supplemental Table 1 for "Tackling the primary healthcare workforce crisis: time to talk about health systems and governance. A comparative assessment of the European region"

**Supplementary material 1**

**Country cases, references and documents**

- **Denmark**

Beich A. Læger vil hellere betjene raske end behandle syge (Information, 13 November 2023:p16). 2023. <https://www.information.dk/anders-beich-0>. Accessed 21 March 2024.

Danske Regioner. Aftale mellem regeringen, Danske Regioner og KL om udmøntning og implementering af en sundhedsreform. 2022. <https://sum.dk/Media/637907113768158579/Aftale%20mellem%20regeringen,%20Danske%20Regioner%20og%20KL%20om%20udm%C3%B8ntning%20og%20implementering%20af%20en%20sundhedsreform.pdf>. Accessed 21 March 2024.

Finansministeriet Trepartsaftale om løn og arbejdsvilkår. Copenhagen: Finansministeriet. Trepartsaftale om løn og arbejdsvilkår. 2023. <https://www.fm.dk>. Accessed 21 March 2024.

Indenrigs- og Sundhedsministerium. Regeringens strategi for coronahåndtering i vintersæsonen 2021/2022. Copenhagen: Indenrigs- og sundhedsministerium. 2021. <https://www.sum.dk>. Accessed 21 March 2024.

Kommissionen for robusthed i sundhedsvæsenet. Robusthedskommissionens anbefalinger. Copenhagen: Sundheds- og Indenrigsministerium; [Robusthedskommissionens anbefalinger – September 2023](https://sum.dk/Media/638336462586551242/Robusthed-Samlet-Rapport-TILG.pdf). 2023. <https://www.sum.dk>. Accessed 21 March 2024.

Kommunernes Landsforening. Udbud af og efterspørgsel efter udvalgte velfærdsuddannelser – 2021. Copenhagen: Kommunernes Landsforening. 2022. Udbud af og efterspørgsel efter udvalgte velfærdsuddannelser. 2021. <https://www.kl.dk>. Accessed 21 March 2024.

Kommunerns Landsforening. Casebank; Casebank – Rekrutteringsinitiativer pa aeldre;- og gesundhedsomradet. 2023. <https://www.kl.dk>. Accessed 21 March 2024.

Lægeforeningen. Arbejdskraftsanalyse 2023. Copenhagen: Lægeforeningen. 2023. <https://laeger.dk/media/fm2fd1oq/arbejdskraftanalyse-2023.pdf>. Accessed 21 March 2024.

Laschke ADL, Blaakaer J, Jensen CF, Larsen MB. Danish general practitioners as gatekeepers for gynaecological patients in regions with different density of resident specailists in gynaecology: in which situations and to whom do hem refer? A cross-sectional study. Sand J Prim Health Care. 2023;41:52–60. <https://doi.org/10.1080/02813432.2023.2165085>

OECD/European Observatory on Health Systems and Policies. Denmark: Country Health Profile 2023, State of Health in the EU. Brussels: OECD/European Observatory on Health Systems and Policies. 2023. <https://doi.org/10.1787/e4f0bee3-en>. Accessed 21 March 2024.

Sundhedsstyrelsen [National Board of Health]. Homepage. 2024. <https://www.sst.dk>. Accessed 21 March 2024.

- **Portugal**

Barros PP; Costa E. Recursos Humanos em Saúde – relatório 2022. Carcavelos: NOVA School of Business and Economics. 2022. <https://www.novasbe.unl.pt/Portals/0/Files/Social%20Equity%20Initiative/Nova%20SBE_KC%20Health_Recursos%20Humanos_2022.pdf?_ga=2.133062733.2146626256.1681719535-2102557819.1677853107&_gl=1*1em6k71*_ga*MjEwMjU1NzgxOS4xNjc3ODUzMTA3*_ga_NRJH2682FN*MTY4MTk4MDA0Mi4xNDAuMS4xNjgxOTgwMTU1LjAuMC4w>. Accessed 22 March 2024.

Biscaia AR, Fehn AC, Pereira A. O médico de família português: uma narrativa. Cadernos De Saúde Pública. 2019;35(1):e00127118.

Biscaia AR, Heleno LC. Primary Health Care Reform in Portugal: Portuguese, modern and innovative. Cien Saude Colet. 2022;22(3):701-712.

Botelho A, Correia I, Fernandes T, Pinto L, Teixeira J, Valente M, et al. Overestimation of health urgency as a cause for emergency services inappropriate use: insights from an exploratory economics experiment in Portugal. Health & Social Care in the Community. 2019;27(4):1031-1041.

Correia T, Gomes I, Nunes P, Dussault, G. Health workforce monitoring in Portugal: Does it support strategic planning and policy-making? Health Policy. 2020;124 (3):303-310.

Lopes D, Castro EA, Simões J. Saúde 2040: Planeamento de Médicos e Enfermeiros em Portugal. Coimbra: Almedina. 2018. <https://novaresearch.unl.pt/en/publications/sa%C3%BAde-2040-planeamento-de-m%C3%A9dicos-e-enfermeiros-em-portugal>. Accessed 22 March 2024.

OECD/European Observatory on Health Systems and Policies. Portugal: Country Health Profile 2023, State of Health in the EU. Paris: OECD. 2023. <https://www.oecd-ilibrary.org/social-issues-migration-health/portugal-country-health-profile-2023_069af7b1-en>. Accessed 21 March 2024.

[Passos Cupertino de Barros](https://onlinelibrary.wiley.com/authored-by/Cupertino+de+Barros/Fernando+Passos) F, [Pereira](https://onlinelibrary.wiley.com/authored-by/Pereira/Filomena) F, [Correia](https://onlinelibrary.wiley.com/authored-by/Correia/Tiago) T, [Ferrinho P.](https://onlinelibrary.wiley.com/authored-by/Ferrinho/Paulo) Primary health care ‘From Alma-Ata to Astana’: Fostering the international debate through the experiences of Portuguese-speaking countries. Int J Health Plann Mgmt. 2022;37(5):2528-2533. <https://doi.org/10.1002/hpm.3511>

Rocha JVM, Telles JL, Heleno B, Rocha THA, da Silva NC, Barbosa P, Santana R. Primary health care reforms in Brazil and Portugal. Public Policy Portuguese Journal. 2020;5(1):40-56.

Temido M, Biscaia A, Correia T, Dussault G, Poole Costa JMF, et al. Melhorar a gestão do SNS. Recursos humanos: o essencial. In Conselho Económico e Social, editors. A Saúde e o Estado: o SNS aos 40 anos. Lisboa: Almedina. 2018, p.187-222. <https://novaresearch.unl.pt/en/publications/melhorar-a-gest%C3%A3o-do-sns-recursos-humanos-o-essencial>. Accessed 22 March 2024.

- **United Kingdom/ England**

Anderson M, O'Neill C, Macleod Clark J, Street A, Woods M, Johnston-Webber C, et al. Securing a sustainable and fit-for-purpose UK health and care workforce. Lancet. 2021;22;397(10288):1992-2011. doi: 10.1016/S0140-6736(21)00231-2.

British Medical Association (BMA). General Medical Services (GMS) contract. London: BMA. 2024. <https://www.bma.org.uk/advice-and-support/gp-practices/gp-contracts/gms-contract>. Accessed 21 March 2024.

Burn E, Fisher R, Locock L, Smith J. A longitudinal qualitative study of the UK general practice workforce experience of COVID-19. Prim Health Care Res Dev. 2022;23:e45. doi: 10.1017/S1463423622000391

Chappell P, Dias A, Bakhai M, Ledger J, Clarke GM. How is primary care access changing? A retrospective, repeated cross-sectional study of patient-initiated demand at general practices in England using a modern access model, 2019-2022. BMJ Open. 2023;13(8):e072944. doi: 10.1136/bmjopen-2023-072944.

DHSC, NHS England. Delivery plan for recovering access to primary care. London: NHS. 2023. <https://www.england.nhs.uk/wp-content/uploads/2023/05/PRN00283-delivery-plan-for-recovering-access-to-primary-care-may-2023.pdf>. Accessed 21 March 2024.

Fisher R, Thorlby R, Alderwick H. Briefing: Understanding primary care networks: Context, benefits and risks. London: Health Foundation. 2019. <https://www.health.org.uk/publications/reports/understanding-primary-care-networks?gclid=Cj0KCQiAwvKtBhDrARIsAJj-kTj0-kLhh_-kF6BRapI0pXCh0OqkyIMqAZ19X0OOa8TYA69nvmrQ1L4aAoRQEALw_wcB>. Accessed 21 March 2024.

Fraser C, Beech J, Mendelsohn E. General practice tracker: Monitoring data on GP appointments and workforce. London: Health Foundation. 2023. <https://www.health.org.uk/news-and-comment/charts-and-infographics/general-practice-tracker>. Accessed 21 March 2024.

Galleta-Williams H, Esmail A, Grigoroglou C, Zghebi S, Zhou A, Hodkinson A, Panagioti M. The importance of teamwork climate for preventing burnout in UK general practice. Eur J Public Health. 2020;30:iv36–iv38. <https://doi.org/10.1093/eurpub/ckaa128>

Karuna C, Palmer V, Scott A, Gunn J. Prevalence of burnout among GPs: a systematic review and meta-analysis. British Journal of General Practice. 2022;72: e316-e324. DOI: 10.3399/BJGP.2021.0441

McDermott I, Spooner S, Goff M, Gibson J, Dalgarno E, Francetic I, et al. Scale, scope and impact of skill mix change in primary care in England: a mixed-methods study. Health Soc Care Deliv Res. 2022;10. Doi: 10.3310/YWTU6690

NHS England. General Medical Services (GMS) contract. London: NHS. 2024. <https://www.england.nhs.uk/gp/investment/gp-contract/>. Accessed 21 March 2024.

NHS England. GP contract reform – investment and evolution. London: NHS. 2024. <https://www.england.nhs.uk/gp/investment/>. Accessed 21 March 2024.

NHS England. NHS Long Term Workforce Plan. London: NHS. 2023. <https://www.england.nhs.uk/long-read/nhs-long-term-workforce-plan-2/>. Accessed 21 March 2024.

NHS England. Primary Care Networks (PCNs) – NHS. London: NHS. 2024. <https://www.england.nhs.uk/primary-care/primary-care-networks/>. Accessed 21 March 2024.

NHS Long Term Plan. NHS Long Term Plan. London: NHS. 2024. <https://www.longtermplan.nhs.uk/>. Accessed 21 March 2024.

Royal College for General Practitioners. Fit for the Future Retaining the GP workforce. London: Royal College of General Practitioners. 2022. <https://www.rcgp.org.uk/getmedia/155e72a9-47b9-4fdd-a322-efc7d2c1deb4/retaining-gp-workforce-report.pdf>. Accessed 21 March 2024.

Shembavnekar N, Buchan J, Bazeer N, Kelly E, Beech J Charlesworth A, et al. REAL Centre Projections: General practice workforce in England. London: Health Foundation. 2022. <https://www.health.org.uk/news-and-comment/charts-and-infographics/general-practice-tracker>. Accessed 21 March 2024.

Zhou AY, Zghebi SS, Hodkinson A, Hann M, Grigoroglou C, Ashcroft DM, et al. Investigating the links between diagnostic uncertainty, emotional exhaustion, and turnover intention in General Practitioners working in the United Kingdom. Front Psychiatry. 2022;13:936067. doi: 10.3389/fpsyt.2022.936067.

- **Germany**

BIBB. Medizinische Fachangestellte. Bonn: BIBB, 2024. [https://www.bibb.de/de/122627.php#](https://www.bibb.de/de/122627.php). Accessed 21 March 2024.

Bundesministerium für Gesundheit (BMG). Ärztliche Behandlung und Versorungsformen, Verbesserung der Versorgung im ländlichen Raum. Berlin: BMG. 2023; <https://www.bundesgesundheitsministerium.de/aerztliche-behandlung>. Accessed 21 March 2024.

European Observatory on Health Systems and Policies (OBS). Germany. Health Systems in Transition. 2021;22(6). <https://eurohealthobservatory.who.int/publications/i/germany-health-system-review-2020>. Accessed 21 March 2024.

Groenewegen PP, Heinemann S, Greß S, Schäfer W. Primary care practice composition in 34 countries. Health Policy. 2015;119:1576–1583. <https://doi.org/10.1016/j.healthpol.2015.08.005>

Institut für hausärztliche Fortbildung im Deutschen Hausärzteverband (IHF). VERAH ist eine hochqualifizierte Weiterbildungsmaßnahme für Medizinische Fachangestellte. Berlin: IHF. 2019. <https://www.verah.de/>. Accessed 21 March 2024.

Kassenärztliche Bundesvereinigung (KBV), AOK Bundesverband, et al. Vertrag über die Hausärztliche Versorgung, DARIS-Archivnr. 1003686213. Berlin: KBV. 2020. <https://www.kbv.de/media/sp/05_Hausaerztl._Versorgung.pdf>. Accessed 21 March 2024.

Kassenärztliche Bundesvereinigung (KBV). Kennzahlen der ambulanten Versorgung auf einen Blick. Berlin: KBV. 2023. <https://www.kbv.de/html/zahlen.php#:~:text=330.000%20Medizinische%20Fachangestellte%20arbeiten%20f%C3%BCr%20niedergelassene%20%C3%84rzte>. Accessed 21 March 2024.

Kringos DS, Boerma WG, Bourgueil Y, Cartier T, Hasvold T, Hutchinson A, et al. The European primary care monitor: structure, process and outcome indicators. BMC Fam Pract. 2010;11:81. <https://doi.org/10.1186/1471-2296-11-81>

Kuhlmann E, Groenewegen PP, Bond C, Burau V, Hunter DJ. Primary care workforce development in Europe: an overview of health system responses and stakeholder views. Health Policy*.* 2018;122(10):1055–1062. <https://doi.org/10.1016/j.healthpol.2018.07.021>

Maier CB, Batenburg R, Birch S, Zander B, Elliott R, Busse R. Health workforce planning: which countries include nurse practitioners and physician assistants and to what effect? Health Policy. 2018;122(10):1085-1092. doi:10.1016/j.healthpol.2018.07.016

OECD. Health statistics. Paris: OECD. 2023. <https://www.oecd.org/health/health-data.htm>. Accessed 21 March 2024.

OECD/European Observatory on Health Systems and Policies. Germany: Country Health Profile 2023, State of Health in the EU. Paris: OECD. 2023. <https://doi.org/10.1787/21dd4679-en>.

Robert-Bosch-Stiftung (RBS). Gesundheitszentren. Wie ein Neustart der Primärversorgung gelingen kann. Stuttgart: RBS. 2021. <https://www.bosch-stiftung.de/sites/default/files/publications/pdf/2021-05/Studie_Primaerversorgung_Gesundheitszentren-fuer-Deutschland.pdf>. Accessed 21 March 2024.

### Robert-Bosch-Stiftung (RBS). Pressemeldung. 2035 fehlen in Deutschland rund 11.000 Hausärzte – Experten empfehlen den Aufbau von Gesundheitszentren. Stuttgart: RBS. 2023. <https://www.bosch-stiftung.de/de/presse/2021/05/2035-fehlen-deutschland-rund-11000-hausaerzte-experten-empfehlen-den-aufbau-von#:~:text=Pressemeldung-,2035%20fehlen%20in%20Deutschland%20rund%2011.000%20Haus%C3%A4rzte%20%E2%80%93%20Experten%20empfehlen%20den,oder%20von%20Unterversorgung%20bedroht%20sein>. Accessed 21 March 2024.

Verband Medizinischer Fachberufe. Pressemitteilung. Krisengipfel der Ärzteschaft soll auch MFA in den Fokus nehmen. 8 January 2024. <https://www.vmf-online.de/verband/presse-news/2024-01-08-krisengipfel>. Accessed 21 March 2024.

World Health Organisation (WHO). Primary care measurement framework and indicators: monitoring health systems through a primary care lens. Geneva: WHO. 2022. <https://www.who.int/news-room/events/detail/2022/02/28/default-calendar/launch-of-the-framework-and-indicators-for-monitoring-primary-health-care>. Accessed 21 March 2024.

- **Netherlands**

Batenburg R, Bosmans M, Versteeg S, Vis E, van Asten B, Vandermeulen L, van der Kruis L. Balans in vraag en aanbod huisartsenzorg. Utrecht: Nivel. 2018. <https://www.nivel.nl/nl/publicatie/balans-vraag-en-aanbod-huisartsenzorg> Accessed 21 March 2024.

Batenburg R, Flinterman L, Vis E, van Schaaik A, de Geit E, Kenes RJ, Dijkers B. Cijfers uit de Nivel-registratie van huisartsen en huisartsenpraktijken. Een acyulaisering voor de periode 2020-2022. Utrecht: Nivel, 2023. <https://www.nivel.nl/sites/default/files/bestanden/1004340.pdf>. Accessed 21 March 2024.

Flinterman L, Groenewegen PP, Verheij R. Zorglandschap en zorggebruik in een veranderende eerste lijn. Utrecht: Nivel, 2018. <https://www.nivel.nl/nl/publicatie/zorglandschap-en-zorggebruik-een-veranderende-eerste-lijn>. Accessed 21 March 2024.

Helderman J-K, Schut F, van der Grinten TED, van de Ven WPMM. Martket-oriented health care reforms and policearning in the Netherlands. Journal of Health Policy, Politics and Law. 2005;30:189-210. DOI:[10.1215/03616878-30-1-2-189](https://doi.org/10.1215/03616878-30-1-2-189)

Kenes RJ, Batenburg R. Cijfers uit de Nivel-registratie van verloskundigen. Utrecht: Nivel. 2021. <https://www.nivel.nl/sites/default/files/bestanden/1004097.pdf>. Accessed 21 March 2024.

Kneepers E. We gaan er alles aan doen om commerciele partijen uit de huisartsenzorg te weren. Strijdvaardigheid maar ook machteloosheid bij de zorgverzekeraar. Medisch Contact. 2023:online. <https://www.medischcontact.nl/actueel/laatste-nieuws/artikel/we-gaan-er-alles-aan-doen-om-commerciele-partijen-uit-de-huisartsenzorg-te-weren>. Accessed 21 March 2024.

Kuhlmann E, Groenewegen PP, Bond C, Burau V, Hunter DJ. Primary care workforce development in Europe: an overview of health system responses and stakeholder views. Health Policy. 2018;122:1055-1062. <https://doi.org/10.1016/j.healthpol.2018.07.021>

LHV. Zorgen om commerciele ketens in de huisartsenzorg. Landelijke Huisartsen Vereniging, 2023; <https://www.lhv.nl/nieuws/zorgen-om-commerciele-ketens-in-de-huisartsenzorg/>

Lindner L, Hayen A. Value-based payment models: an assessment of the Menzis Shared Savings programme in the Netherlands. DELSA/HEA/WD/HWP(2023)10. Paris: OECD. 2023. <https://dx.doi.org/10.1787/0810f2ba-en>. Accessed 20 March 2024.

Maier CB, Batenburg R, Birch S, Zander B, Elliott R, Busse R. Health workforce planning: which countries include nurse practitioners nad pyhsician assistants and to what effect? Health Policy. 2018;122:1085-1092.

OECD/European Observatory on Health Systems and Policies. Netherlands: Country Health Profile 2023, State of Health in the EU. Paris: OECD. 2023. <https://doi.org/10.1787/3110840c-en>. Accessed 21 March 2024.

Rafferty AM, Busse R, Zandre-Jentsch B, Sermeus W, Bryneel L. Strengthening health systesm through nursing. Evidence from 14 European countries. Copenhagen: WHO/ Europan Observatory on Helath Systems and Policies. 2019. <https://www.ncbi.nlm.nih.gov/books/NBK545724/pdf/Bookshelf_NBK545724.pdf> Accessed 20 March 2024.

Schuurmans J, Stalenhoef H, Bal R, Wallenburg I. All the good care: valuation and task differentiation in older person care. Sociology of Health & Illness*.* 2023;45:1560–1577. <https://doi.org/10.1111/1467-9566.13654>.

Schuurmans J, van der Woerd O, Bal R, Wallenburg I. Regionalisering in de ouderenzorg: een beleidssociologisch perspectief op grootschalige verandering. Beleid & Maatschappij*.* 2022;49:220-239. DOI:[10.5553/BenM/138900692022005001](http://dx.doi.org/10.5553/BenM/138900692022005001)

Veldkamp R, Meijer W. Zorg door de Fysiotherapeut. Nivel Zorgregistraties Eerste Lijn: jaarcijfers 2022 en trendcijfers 2018-2022. Utrecht: Nivel. 2023. <https://www.nivel.nl/nl/publicatie/zorg-door-de-fysiotherapeut-nivel-zorgregistraties-eerste-lijn-jaarcijfers-2021-en>. Accessed 21 March 2024.

Wallenburg I, Putters K, van der Grinten TED. Scheiden of verbinden. Dilemma’s voor beleid en uitvoering in zorg en dienstverlening. Beleid & Maatschappij. 2023;50:64-72. DOI:[10.5553/BenM/138900692023050001008](http://dx.doi.org/10.5553/BenM/138900692023050001008)

- **Switzerland**

Blick.ch. Den Hausärzten laufen die Medizinischen Praxisassistentinnen davon. Blick.ch, 13 October 2023. <https://www.blick.ch/schweiz/westschweiz/wallis/wenig-lohn-viel-stress-den-hausaerzten-laufen-die-praxisassistentinnen-davon-id19035620.html>. Accessed 21 March 2024.

Bundesamt für Gesundheit (BAG). Die gesundheitspolitische Strategie des Bundesrates 2020–2030. Liebefeld: Bundesamt für Gesundheit. 2019. <https://www.bag.admin.ch/dam/bag/de/dokumente/nat-gesundheitsstrategien/gesundheit-2030/strategie-gesundheit2030.pdf.download.pdf/strategie-gesundheit-2030.pdf>. Accessed 21 March 2024.

Bundesamt für Gesundheit (BAG). Höchstzahlen für Ärztinnen und Ärzte. Liebefeld: Bundesamt für Gesundheit. 2023. <https://www.bag.admin.ch/bag/de/home/versicherungen/krankenversicherung/leistungserbringer/hoechstzahlen-aerzte-aerztinnen.html>. Accessed 21 March 2024.

Bundesamt für Gesundheit (BAG). Umsetzung Pflegeinitiative (Artikel 117b BV). Liebefeld: Bundesamt für Gesundheit. 2023. <https://www.bag.admin.ch/bag/de/home/berufe-im-gesundheitswesen/gesundheitsberufe-der-tertiaerstufe/vi-pflegeinitiative.html>. Accessed 21 March 2024.

Bundesamt für Gesundheit (BAG). Gesundheitsberuferegister GesReg. Liebefeld: Bundesamt für Gesundheit. 2022. <https://www.bag.admin.ch/bag/de/home/berufe-im-gesundheitswesen/gesundheitsberufe-der-tertiaerstufe/gesundheitsberuferegister.html>. Accessed 21 March 2024.

Carron T, Domeisen Benedetti F, Fringer, A, Fierz K, Petreymann-Bridevaux I. Integrated care models in Swiss primary care: An embedded multiple case study. J Eval Clin Pract. 2023;29:1025-1038. <https://doi-org/10.1111/jep.13891>

Cartier T, Senn N, Cornuz J, Bourgueil Y. Switzerland. In: Kringos DS, Boerma WGW, Hutchinson A, Saltman R, editors. Building primary care in a changing Europe: Case studies. Observatory Studies Series, No. 40. Copenhagen: European Observatory on Health Systems and Policies. 2015; chapter 29. <https://eurohealthobservatory.who.int/publications/i/building-primary-care-in-a-changing-europe-study>. Accessed 21 March 2024.

De Pietro C, Camenzind P, Sturny I, Crivelli L, Edwards-Garavoglia S, Spranger A, et al. Switzerland: Health System Review. Health Syst Transit. 2015;17(4):1-288.

Federal Office of Public Health. Statistics of medical professions 2022. Liebefeld: Federal Office. 2023. <https://www.bag.admin.ch/dam/bag/de/dokumente/berufe-gesundheitswesen/medizinalberufe/statistiken/medizinalberufestatistik_engl_2022.pdf.download.pdf/Statistics%20of%20medical%20professions%202022.pdf>. Accessed 21 March 2024.

Federal Department of Home Affairs (FDHA). Swiss Health Foreign Policy 2019–2024. Bern: FDHA. 2019. <https://www.bag.admin.ch/bag/en/home/strategie-und-politik/internationale-beziehungen/schweizer-gesundheitsaussenpolitik.html>. Accessed 21 March 2024.

Groenewegen P, Heinemann S, Greß S, Schäfer W. Primary care practice composition in 34 countries. Health Policy. 2015;119(12):1576-83. <https://doi.org/10.1016/j.healthpol.2015.08.005>.

Josi, R., De Pietro, C. Skill mix in Swiss primary care group practices – a nationwide online survey. BMC Fam Pract. 2019;20:39. <https://doi.org/10.1186/s12875-019-0926-7>

Lobsiger M, Liechti D. Berufsaustritte und Bestand von Gesundheitspersonal in der Schweiz. Eine Analyse auf Basis der Strukturerhebungen 2016–2018 (Obsan Bericht 01/2021). Neuchâtel: Schweizerisches Gesundheitsobservatorium. 2021; p6. file:///C:/Users/asus/Downloads/7.3_obsan_01_2021_bericht_berufsaustritte_GP.pdf. Accessed 21 March 2024.

Obsan Indicators. Expenditure on prevention and health promotion by financing scheme. Schweizerisches Gesundheitsobservatorium (Obsan). 2023. <https://ind.obsan.admin.ch/en/indicator/monam/expenditure-on-prevention-and-health-promotion-by-financing-scheme>. Accessed 21 March 2024.

OECD. Health at a Glance 2021: Health expenditure on primary health care. Paris: OECD. 2021. <https://www.oecd-ilibrary.org/sites/0bb3b147-en/index.html?itemId=/content/component/0bb3b147-en>. Accessed 21 March 2024.

OECD.stat. Healthcare Resources. Paris: OECD. 2023. [https://stats.oecd.org/Index.aspx?ThemeTreeId=9#](https://stats.oecd.org/Index.aspx?ThemeTreeId=9). Accessed 21 March 2024.

Pahud O. Ärztinnen und Ärzte in der Grundversorgung – Situation in der Schweiz und im internationalen Vergleich. Analyse des International Health Policy (IHP) Survey 2019 der amerikanischen Stiftung Commonwealth Funds. Schweizerisches Gesundheitsobservatorium (Obsan). 2019. <https://www.obsan.admin.ch/de/publikationen/2019-aerztinnen-und-aerzte-der-grundversorgung-situation-der-schweiz-und-im>. Accessed 21 March 2024.

PhysioSwiss. Direktzugang: Faktencheck der Antwort des Bundesrats. PhysioSwiss. 2023.: <https://www.physioswiss.ch/de/news/2023/direktzugang-faktencheck>. Accessed 21 March 2024.

Santésuisse. Statistique des assurés date de début du traitement 2008. Pool de données. Solothurn: Santésuisse. 2009. <http://www.santesuisse.ch/datasheets/files/200909221629160.pdf>. Accessed 21 March 2024.

Stierli R, Rozsnyai Z, Felber R, Jörg R, Kraft E, Exadaktylos AK, Streit S. Primary Care Physician Workforce 2020 to 2025 – a cross-sectional study for the Canton of Bern. Swiss Med Weekly. 2021;151(3536):w30024. <https://smw.ch/index.php/smw/article/view/3062>. Accessed 21 March 2024.

Swiss Confederation. Switzerland implements the 2030 Agenda for Sustainable Development. Switzerland’s Country Report 2018. Swiss Confederation, 2018; <https://regiosuisse.ch/sites/default/files/2018-06/laenderbericht-der-schweiz-2018_EN.pdf>

Tandjung R, Morell S, Hanhart A, Haefeli A, Valeri F, Rosemann T, Senn O. Referral determinants in Swiss primary care with a special focus on managed care. PLoS One. 2017;12: e0186307. <https://doi.org/10.1371/journal.pone.0186307>

The Commonwealth Fund. Overworked and Undervalued: Unmasking Primary Care Physicians’ Dissatisfaction in 10 High-Income Countries. London: The Commonwealth Fund. 2023. <https://www.commonwealthfund.org/publications/issue-briefs/2023/aug/overworked-undervalued-primary-care-physicians-10-countries>. Accessed 21 March 2024.

Zeller A, Giezendanner S, Resultate der 4. Workforce Studie 2020. Swiss Health Web. 2020. <https://www.swisshealthweb.ch/de/article/doi/phc-d.2020.10311>. Accessed 21 March 2024.

- **Kazakhstan**

Abzaliyeva A, Kausova G, Abdraimova E, Ismagilova A, Mamyrbekova S. Availability of general practice workforce and basic health indicators in the Republic of Kazakhstan: 2015-2019. Maced J Med Sci. 2022;10(E):452-7. <https://oamjms.eu/index.php/mjms/article/view/7880>. Accessed 21 March 2024.

Eriksen A, Litvinova Y, Rechel B. Health Systems in Action: Kazakhstan. Copenhagen: WHO. 2022. <https://eurohealthobservatory.who.int/publications/i/health-systems-in-action-kazakhstan-2022>. Accessed 21 March 2024.

Katsaga A, Kulzhanov M, Karanikolos M, Rechel B. Kazakhstan: Health System Review. Health Systems in Transition. 2012;14(4):1-154. <https://pubmed.ncbi.nlm.nih.gov/22894852/>. Accessed 21 March 2024.

Koichubekov B, Kharin A, Sorokina M, Korshukov I, Omarkulov B. System dynamics modelling for general practitioner workforce forecasting in Kazakhstan. Ann Ig. 2021;33(3):242-253. <https://doi.org/10.7416/ai.2020.2391>

OECD. OECD Reviews of Health Systems: Kazakhstan 2018. Paris: OECD. 2018. <https://doi.org/10.1787/9789264289062-en>. Accessed 21 March 2024.

Rechel B, Sydykova A, Moldoisaeva S, Sodiqova D, Spatayev Y, Ahmedov M, et al. Primary care reforms in Central Asia – On the path to universal health coverage? Health Policy Open. 2023;5:100110. <https://doi.org/10.1016/j.hpopen.2023.100110>.

WHO Regional Office for Europe. Transformation of primary health care in Kazakhstan: moving towards a multidisciplinary model. Primary health care policy paper series. Copenhagen: WHO. 2023. <https://www.who.int/europe/publications/i/item/WHO-EURO-2023-8269-48041-71196>. Accessed 21 March 2024.

WHO Regional Office for Europe. Kazakhstan: multidisciplinary teams for better alignment of primary health care services to meet the needs and expectations of people. Copenhagen: WHO. 2022. <https://www.who.int/andorra/publications/m/item/kazakhstan-multidisciplinary-teams-for-better-alignment-of-primary-health-care-services-to-meet-the-needs-and-expectations-of-people-(2021)>. Accessed 21 March 2024.

WHO. Kazakhstan gears up to launch social health insurance. Bull World Health Organ. 2016;94(11):792-793. <https://doi.org/10.2471/BLT.16.031116>

- **Romania**

Coman M-A, Nemeș D, Gati G, Paina L, Brînzac M-G, Ungureanu MI. Building the primary healthcare workforce in Romania to promote child and adolescent: National overview and county case-study. Babes-Bolyaj University: Romania. 2022. <https://www.unicef.org/romania/media/9556/file/Building%20the%20primary%20healthcare%20workforce%20in%20Romania%20to%20promote%20child%20and%20adolescent:%20National%20overview%20and%20county%20case-study.pdf>. Accessed 21 March 2024.

Coman M-A, Nemeș D, Gati G, Paina L, Brînzac M-G, Ungureanu MI. Building the primary healthcare workforce in Romania to promote child and adolescent well-being. Babes-Bolyaj University: Romania. 2022. <https://www.unicef.org/romania/media/9546/file/Building%20the%20primary%20healthcare%20workforce%20in%20Romania%20to%20promote%20child%20and%20adolescent%20well-being.pdf>. Accessed 21 March 2024.

Duran A, Chanturidze T, Gheorghe A, Moreno A. Assessment of Public Hospital Governance in Romania: Lessons From 10 Case Studies. Int J Health Policy Manag, 2019;8:199. <https://doi.org/10.15171/IJHPM.2018.120>.

Frone S. Progress and prospects of some Sustainable Development Goals in Romania. Institute of National Economy: Bucharest. 2020. <https://doi.org/10.2478/9788366675261-014>.

Jullien S, Mateescu I, Brînzac M-G, Dobocan C, Pop I, Weber MW, et al. Unnecessary hospitalisations and polypharmacy practices in Romania: A health system evaluation for strengthening primary health care. Journal of Global Health. 2023;13: 4039. <https://doi.org/10.7189/JOGH.13.04039>

Kringos DS, Boerma WGW, Hutchinson A, Saltman R, editors. Building primary care in a changing Europe: Case studies. Observatory Studies Series, No. 40. Copenhagen: WHO. 2015. <https://www.ncbi.nlm.nih.gov/books/NBK459010/>. Accessed 21 March 2024.

OECD.stat. Healthcare Resources. Paris: OECD. 2023. [https://stats.oecd.org/Index.aspx?ThemeTreeId=9#](https://stats.oecd.org/Index.aspx?ThemeTreeId=9). Accessed 21 March 2024.

OECD/European Observatory on Health Systems and Policies. *Romania: Country Health Profile 2023*, State of Health in the EU. Paris: OECD. 2023. <https://doi.org/10.1787/f478769b-en>. Accessed 21 March 2024.

Petre I, Barna F, Gurgus D, Tomescu LC, Apostol A, Petre I, et al. Analysis of the healthcare system in Romania: a bief review. Healthcare. 2023;11(14):2069. <https://doi.org/10.3390/healthcare11142069>.

Suciu ŞM, Popescu CA, Ciumageanu MD, Buzoianu AD. Physician migration at its roots: a study on the emigration preferences and plans among medical students in Romania. Hum Resour Health 2017;15:6. <https://doi.org/10.1186/s12960-017-0181-8>.

Vlădescu C, Scîntee SG, Olsavszky V, Hernández-Quevedo C, Sagan A. Romania: health systems in transition. Health Systems in Transition. 2016;18:4. <https://eurohealthobservatory.who.int/publications/i/romania-health-system-review-2016>. Accessed 21 March 2024.

Wang H, Chukwuma A, Comsa R, Dmytraczenko T, Gong E, Onofrei L. Generating political priority for primary health care reform in Romania. Health Systems & Reform. 2021;7:2. <https://doi.org/10.1080/23288604.2021.1898187>.

- **Serbia**

AHEAD National policy dialogue: Serbia – policy options for addressing medical deserts. 2023. <https://ahead.health/wp-content/uploads/2023/04/SER-AHEAD_-Serbia-policy-brief_English.pdf>. Accessed 21 March 2024.

Bjegovic-Mikanovic V, Vasic M, Vukovic D, Jankovic J, Jovic-Vranes A, Santric-Milicevic M, et al. Serbia: Health System Review. Health Systems in Transition. 2019;21:3. <https://eurohealthobservatory.who.int/publications/i/serbia-health-system-review-2019>. Accessed 21 March 2024.

Đukanović K, Bogdanović Vasić S. Palijativna nega kao integralni deo zdravstvene nege. Sestrinska reč. 2020;23(80):11-14. <https://scindeks.ceon.rs/article.aspx?artid=0354-84222080011Q>. Accessed 21 March 2024.

Eurostat Data. Data. Brussels: European Union. 2023. <https://ec.europa.eu/eurostat/data/database>. Accessed 21 March 2024.

Health Care Law, "Official Gazette of RS", no. 25/2019 and 92/2023. (unpublished information)

Health Insurance Law, “Official Gazette of RS”, no. 25/2019 and 92/2023. (unpublished information)

Institute of Public Health of Serbia. Health Statistical Yearbook of Republic of Serbia 2022. Belgrade: Institute of Public Health of Serbia. 2023; p34. <https://www.batut.org.rs/index.php?lang=2>. Accessed 22 March 2024.

Milošević M, Topalović M, Jović-Vraneš A. Scope of preventive services provided by chosen doctors in primary health care, in the Republic of Serbia, in the period between 2013 and 2017. Serbian Journal of the Medical Chamber. 2022;3(3):300-316. <https://doi.org/10.5937/smclk3-39707>.

Ministry of Health of the Republic of Serbia. The Midterm Health Action Plan of the Republic of Serbia (2022-2025). Belgrade: Ministry of Health. 2022. <https://www.zdravlje.gov.rs/view_file.php?file_id=2624&cache=sr>. Accessed 21 March 2024.

National Employment Service of the Republic of Serbia. Unemployment, requests for job-matching services in vacancy filling by occupations groups and sex during November 2023. Monthly Statistical Bulletin number 255. Belgrade: National Employment Service of the Republic of Serbia. 2023; p16; <https://www.nsz.gov.rs/filemanager/Files/Dokumenta/Statisti%C4%8Dki%20bilteni/2023/Bilten%20NSZ%20-%20Novembar%202023.pdf>. Accessed 21 March 2024.

National Human Development Report – Serbia 2022. Human Development in Response to Demographic Change. UNDP Serbia. 2022. <https://hdr.undp.org.rs/wp-content/uploads/2023/05/National-Human-Development-Report-Serbia-2022-e1.pdf>. Accessed 21 March 2024.

Regulation on the Plan of the Network of Health Institutions. Official Gazette of the RS, no. 5/2020, 11/2020, 52/2020, 88/2020, 62/2021, 69/2021, 74/2021 and 95/2021. [Uredba o planu mreže zdravstvenih ustanova]. <https://www.paragraf.rs/propisi/uredba_o_planu_mreze_zdravstvenih_ustanova.html>. Accessed 21 March 2024.

Richardson E, Bjegovic-Mikanovic V. Health systems in action: Serbia. Brussels: European Observatory on Health Systems and Policies. 2022. <https://eurohealthobservatory.who.int/publications/i/health-systems-in-action-serbia-2022>. Accessed 21 March 2024.

Santric Milicevic M, Vasic M, Edwards M. Mapping the governance of human resources for health in Serbia. Health Policy. 2015;119:1613–1620. <https://doi.org/10.1016/j.healthpol.2015.08.016>

Statistical Office of the Republic of Serbia. Progress report on the implementation of sustainable development goals by 2030 in the Republic of Serbia, report for 2022. Belgrade: Statistical Office of the Republic of Serbia. 2023. file:///C:/Users/WINDOWS%2010/Downloads/https___sdgs4all.rs_wp-content_uploads_2023_05_SDG_Izvestaj_2022.pdf. Accessed 21 March 2024.

Statistical Office of the Republic of Serbia. Statistical Yearbook. Belgrade: Statistical Office of the Republic of Serbia. 2023; p68. <https://www.stat.gov.rs/en-US/publikacije/>. Accessed 21 March 2024.

WHO. Global Health Expenditure Database. Health expenditure profile: Serbia. Geneva: WHO. 2023. <https://apps.who.int/nha/database/country_profile/Index/en>. Accessed 21 March 2024.
