## Supplemental Table 2 for "Tackling the primary healthcare workforce crisis: time to talk about health systems and governance. A comparative assessment of the European region"

**Supplementary material 2**

**Matrix**

| Country |
| --- |
| PHC sector overview |
| - Provision |
| - Finance |
| - Governance |
| PHC healthcare workforce |
| - Main professions/occupations |
| - Education |
| - Labour market figures |
| - HCWF data, monitoring, planning |
| - HCWF governance |
| PHC workforce policy |
| - Problems/challenges |
| - Policy |
| - Interventions/policy implementation |
| Global goals/frameworks |
| - Role of the WHO PHC concept |
| - Role of the SDGs |

Background information

References, documents
